# Gene therapy using optimized LentiHBB^T87Q^ vector in two patients with transfusion dependent β-thalassemia

**DOI:** 10.1101/2023.03.21.23287513

**Authors:** Nan Han, Yue Li, Wenjie Ouyang, Guoyi Dong, Honglian Guo, Yue Chen, Yan Huang, Xinru Zeng, Huilin Zou, Jiajun He, Wenwen Yao, Chao Liu, Sixi Liu

## Abstract

**Background:** Gene therapy is gradually becoming recognized as a possibly curative therapeutic strategy for transfusion-dependent β-thalassemia (TDT). Gene therapy addresses the problem of donor scarcity through the application of autologous hematopoietic stem cells (HSCs), which also can reduce the risks that accompany allogeneic HSC transplantation. When using gene addition strategy, lentiviral vector is critical for the efficacy and safety of β-thalassemia gene therapy. In our preclinical studies, LentiHBB^T87Q^ vector with optimized backbone was developed to efficiently restore β–globin expression in HSCs-derived erythroblasts of TDT patients with minimal risk of tumorigenesis. Here, we presented the clinical trial results of gene therapy using LentiHBB^T87Q^ vector in two TDT patients.

**Method:** In an ongoing phase 1/2 trial (NCT05745532), auto-HSCs were mobilized from two TDT patients, and then transduced with LentiHBB^T87Q^ vector. The gene-modified auto-HSCs is called HGI-001 injection. After four-day consecutive myeloablative conditioning, these two patients were administrated with HGI-001 injection via intravenous infusion. Medical examinations were performed in the transplantation unit to monitor patients’ status till the patients were clinically stable. Then, 24-month following-up visits are conducted to assess the safety and efficacy of HGI-001 injection. The safety endpoints of this clinical study include the incident and severity of adverse events (AEs); transplant-related mortality or disability events within 100 days post drug product infusion; vector-related replication competent lentivirus (RCL) and clonal variations containing specific viral integration sites; overall survival during this clinical trial. The major efficacy endpoint is the percentage of subjects with average vector copy number (VCN) in peripheral blood mononuclear cells (PBMCs) >0.1, and average expression of exogenous HbA^T87Q^ >2.0g/dL at the 24^th^ month after reinfusion of HGI-001 injection

**Results:** The rapid neutrophil and platelet engraftment successfully happened after reinfusion of HGI-001 injection. The two patients with non-β^0^/β^0^ genotype have been transfusion-independent for 24 months and 21 months post-treatment. At the last visit, the levels of HbA^T87Q^ are 7.3 and 6.9g/dL, and the levels of total hemoglobin are 9.8 and 10.1 g/dL. After the two subjects stopped transfusions, the iron overload has been alleviated without iron chelation treatment. Most AEs are myeloablative conditioning related, and can be controlled through clinically standard therapeutic managements. No clone dominance related to vector integration nor RCL has been observed.

**Conclusion:** Gene therapy with optimized LentiHBB^T87Q^ vector (HGI-001 injection) assist two TDT patients become transfusion-independent without serious adverse events related to the product.

## Introduction

Thalassemia, an anemia caused by disorders of hemoglobin production, is regarded as the most widespread monogenic hereditary disease worldwide^1^. In addition to being prevalent in the Mediterranean region, South-East Asia, the Indian subcontinent, and the Middle East^2^, β-thalassemia has become a worldwide challenge that has spread to most of Europe, the Americas, and Australia because of migration^3^. According to a report from World Health Organization in 2008, there could be roughly 25,500 infants born in the world each year, who have transfusion-dependent β-thalassemia (TDT)^4^. The etiology of β-thalassemia is due to various mutations in the β-globin gene, and more than 350 mutations have been described^5^. These mutations are generally categorized as β^+^ or β^0^ to describe the severity of thalassemia, with β^+^ being a mild mutation resulting in relatively decreased β-globin synthesis and β^0^ being a severe mutation resulting in the absolute non-production of β-globin chains. Hemoglobin E (Hb E) is a common β-globin variation that has a mild form of β- thalassemia, as its slightly lower rate of production and instability with α-globin under oxidant stress. Because of the coinheritance of Hb E and β^0^, Hb E/β^0^ occurs with an extremely high frequency, accounting for approximately half of all clinical patients of β-thalassemia major globally^6,7^. In β-thalassemia, the disequilibrium of α- globin and β-globin chains (excess free α-globin and insufficient β-globin proteins) is mainly involved in ineffective erythropoiesis and chronic hemolysis^8,9^, which together result in microcytic hypochromic anemia^10^. To compensate the loss of β-globin, an increasing of fetal hemoglobin (Hb F) level is observed in the several various cohorts of β-thalassemia patients ^11-13^.

Nevertheless, TDT patients require long-term red-cell transfusion for survival and prophylaxis of various complications for survival^1,14^. Allogeneic hematopoietic stem-cell transplantation (HSCT) using an HLA-matched sibling donor (MSD) is currently the only available cure for TDT patients ^5,15^. However, HSCT as a curative therapy is limited by insufficient HLA-MSDs and by the risk of graft-versus-host disease, graft failure and long-term complications in real life^16,17^. Therefore, gene therapy through autologous transplantation of genetically modified hematopoietic stem cells (HSCs) represents an innovative therapeutic prospect to treat thalassemia, and may fulfill the unmet medical need in the field. Notably, an ex-vivo lentiviral vector gene therapy, Bluebird Bio’s ZYNTEGLO, is recently approved by FDA for the treatment of β-thalassemia in adult and pediatric patients with TDT. In its ongoing Phase 3 trials (HGB-207), 91% subjects (20 of 22) with non-β^0^/β^0^ genotype achieved transfusion independence after gene therapy with BB305 vector^18^. In another two clinical trials, blood transfusions were continued in 4 out of 7 and 4 out of 4 TDT subjects who had received autologous CD34^+^ cells transduced with GLOBE vector and TNS9.3.55 vector respectively^19,20^. These results demonstrate the importance of lentiviral vector, lentiviral transduction efficiency as well as the manufacturing procedure^21^. We previously developed a lentiviral vector with optimized backbone named LentiHBB^T87Q^, containing an exogenous T87Q β-globin gene (HBB^T87Q^) driven by a specific reconstituted locus control region. In preclinical studies, LentiHBB^T87Q^ can efficiently express exogenous *HBB* mRNA and restore HBB protein in HSCs-derived erythroblasts of TDT patients with minimal risk of tumorigenesis^22^. In addition, clinical-grade LentiHBB^T87Q^ was manufactured and lentiviral transduction procedure was optimized at a large scale which is applicable to clinical trial^22^. The auto-HSCs transduced by LentiHBB^T87Q^ are named HGI-001 injection.

We present the results from an ongoing phase 1/2 clinical trial that evaluate the safety and efficacy of gene therapy for β-thalassemia using the LentiHBB^T87Q^ vector. In this study, five TDT patients already received HGI-001 injection and were followed up for 2 to 24 months. Here, we report the results of only two patients with TDT with a follow-up period of more than 21 and 24 months.

## Methods

### Study design and Participants

This single center, open-label, single-arm, phase 1/2, pilot trial (NCT05745532) was launched in 2020 to evaluate the safety and efficacy of HGI-001 injection. The protocol was reviewed and approved by the Medical Ethics Committee of Shenzhen Children’s Hospital. The study was conducted in complete adherence to the Declaration of Helsinki.

The pediatric patients were enrolled at the Hematology & Oncology of Shenzhen Children’s Hospital (Shenzhen, China). Subjects were pediatric patients aged from 8 to 16 years and diagnosed with TDT, regardless of β-thalassemia genotypes. Here, the standard of TDT was that participants who annually received ≥100ml packed red-blood cells per kilogram of body weight, or ≥ 8 times transfusion each year for at least 2 years under regular β-thalassemic management^23^. The eligibility criteria in our study are as following: parents or guardians of subjects signed informed consent, and the patient gave assent before screening; regular blood transfusion for at least 3 months prior to screening (Hb≥9.0g/dL); serum ferritin<3000μg/L; cardiac magnetic resonance imaging (MRI) T2* >10ms and liver MRI T2* >1.4ms, being clinically stable and eligible to undergo autologous HSCT.

Patients were excluded if they had a known and available human leukocyte antigen (HLA)-matched family/unrelated donor, prior receipt of HSCT and gene therapy, a history of splenectomy, uncorrected bleeding or seizure disorder, pulmonary hypertension without effective intervention, positive for blood cell antibody in antibody screening test, clinically significant and active bacterial, viral, fungal, or parasitic infection, any prior or current malignancy, myeloproliferative, immunodeficiency disorder or autoimmune disease, other medical conditions who are not eligible to participate in the study (such as advanced liver disease, kidney disease or cardiac disease, etc.). Complete inclusion and exclusion criteria are shown in Table S1 in the Supplementary Appendix.

Inclusion and exclusion were evaluated based on the patient’s medical history, physical examination, complete blood count, iron studies, liver function, and serum biochemistry, coagulation tests, serology tests, bone marrow biopsy, immunological testing, red blood cell antibody screen, imaging assessment, pulmonary function tests and so on. All the medical examinations should be finished within 28 days. Subjects who had used Hydroxyurea, Ruxolitinib, Decitabine or Cytarabine in the 3 months prior to screening did not meet the inclusion criteria. Patients who had enrolled in an investigational drug within the previous 4 weeks were excluded were not included.

### Procedures of Gene therapy

Our study mainly includes the processes of screening, mobilization and apheresis, HGI-001 injection manufacturing, myeloablative conditioning, reinfusion of HGI-001 injection, and following-up for 24 months (Figure S1). Before gene therapy, patients were confirmed eligible for the trial following the complete evaluations and examinations at their screening phase. Hemoglobin (Hb) level of each patient was recommended to preserve at ≥9.0g/dL for at least three months prior to hematopoietic stem and progenitor cell mobilization. The combination of G-CSF and plerixafor was used for HSCs mobilization, and then cells were collected by apheresis under the monitoring of CD34^+^ cell count. Patients’ first apheresis would be performed on the 5^th^ day of mobilization cycle (the day after plerixafor administration) (Figure S2). The harvested HSCs were immediately sent to certified GMP facility (Shenzhen, China) for HGI-001 injection manufacture, including isolation of CD34^+^ cells, transduction with LentiHBB^T87Q^, and then cryopreservation with programmed cooling and stored in gaseous liquid nitrogen. Before re-infusion, HGI injection should pass the final quality inspection release. Meanwhile, at least 2 million unmanipulated CD34^+^ cells/kilogram per patient were preserved as backup cells for rescuing in case of engraftment failure.

When the indicated patient’s drug product (HGI-001 injection) was ready for clinical use and stored at the clinical site, the patient was medically examined and assessed to reconfirm that the myeloablative conditioning could be performed. The patient’s myeloablative conditioning required busulfan for four consecutive days at a start dose of 3.2mg/kg/day. To reduce the risks of adverse events (AEs) caused by busulfan, it is recommended to administer the busulfan every 6 hours (q6h), divided into four doses of 0.8/mg/kg/day, with each time lasting approximately for two hours. The dose for the remaining 3 days was adjusted according to the first dose pharmacokinetics to achieve the target concentration. It is recommended that the target area under curve (AUC) of every 6 hours (q6h) dosing is 1050 (range 950 to 1125) μM*min. After a minimum 48-hour washout, the HGI-001 injection was administrated to the patient by intravenous reinfusion at the clinical site (Figure S3). Prior to reinfusion, the HGI-001 injection must be thawed at 37°C with care taken to avoid contamination, and the reinfusion should be completed within 30 minutes after cell thawing. The minimum dose of CD34^+^ cells were above 3.0 million/kg, and the day of HGI-001 reinfusion was defined as Day 0. Patients were followed daily in the transplant unit for AEs, and medical examinations were conducted to monitor bone marrow engraftment. Once the patients’ status achieved medical stability, patients were allowed to discharge from transplant units, and then followed in this protocol for a period of 24 months after reinfusion.

### The endpoints of safety and efficacy

The safety endpoints of this study include the incident and severity of clinical adverse events during the trial; transplant-related mortality or disability events within 100 days post drug product infusion; vector-related replication competent lentivirus (RCL) and clonal variations containing specific viral integration sites; overall survival during this clinical trial. AEs (including severe adverse events) were recorded from the time informed consent was signed and monitored throughout an entire trial up to 24 months after HGI-001 infusion. In addition, the clonal abundance exceeding 20% for more than 3 months is defined as clonal dominance through peripheral blood integration site analysis (ISA) after gene therapy.

The primary efficacy endpoint is to obtained the percentage of subjects with average vector copy number (VCN) in peripheral blood mononuclear cells (PBMCs) >0.1, and average expression of exogenous HbA^T87Q^ >2.0g/dL at the 24^th^ month after reinfusion of HGI-001 injection. The secondary efficacy endpoints are as follow: (1) the percent of subjects with average VCN in PBMCs >0.1, and expression of exogenous HbA^T87Q^ >2.0g/dL at 12 months after reinfusion of HGI-001 injection; (2) percent of subjects with successful bone marrow engraftment (criteria for successful engraftment: absolute neutrophil count>0.5×10^9^/L for 3 consecutive days; and platelet count is maintained at more than 20 ×10^9^/L for 7 consecutive days without supportive platelet transfusion); (3) the reduction in frequency and volume of red-cell transfusions compared with the average frequency and volume of transfusions in the 12 months prior to enrollment; (4) transfusion independent (TI), which is indicated that subjects stop to receive red-cell transfusion for minimum 12 consecutive months, while maintaining weight average Hb level of 9.0g/dL or higher; (5) transfusion-free survival, as defined by the duration of time a subject has met TI criteria and remained transfusion-free; (6) persistence of VCN and HbA^T87Q^ expression in PBMCs and bone marrow throughout the study. Additionally, exploratory endpoints are the assessment of iron metabolism, hemolysis, and erythropoiesis via relevant clinical laboratory tests.

The pharmacodynamic endpoints (as called Cytokinetics) was to evaluate exogenous globin expression induced by HGI-001 injection based on two parameters, including changes in VCN of PBMCs and proportion of HbA^T87Q^ in total hemoglobin over time.

### Statistical Analysis

The treated population, including all patients who received the drug product (HGI-001 injection), was the primary population for this analysis. The data from each patient were analyzed separately. For continuous variables, median or average, minimum, and maximum values are exhibited. The method of ISA is shown in the Supplementary Appendix. Due to the CoVID-19 pandemic, data are missing at some time points. The 15M visit was missing and the follow-up 12M was migrated to 14M for F001. For F002, the visit of 9M was missing and 18M visit was rescheduled to 19M.

### Results

### Basic information of patients

On June 2020, two subjects (F001 and F002) formally recruited into the study and signed the informed consent. The age of F001 and F002 were enrolled at the age met the present inclusion criteria of our study, and the course of β-thalassemia major for F001 and F002 was more than 5 years. The genotypes of two patients were β^0^/β^+^ and β^E^/β^+^ respectively, and the total red-cell transfusion volume was 332.6 and 257.7ml/kilogram in the 12 months before enrollment. Pre-therapy baseline serum ferritin level was defined as the value prior to the mobilization. Neither of them had severe cardiac or liver iron overload by investigators’ evaluation, and they were managed under regular packaged red-cell transfusions and iron chelation treatment before mobilization and apheresis. The complete basic information of two patients is in Table 1.

**Table 1.** The basic demographic and medical information

|  | **F001** | **F002** |
| --- | --- | --- |
| **Basic information** |  |  |
| Sex | Male | Female |
| Race | Asian | Asian |
| Height(cm) | 116 | 145 |
| Weight(kg) | 23 | 39 |
| **History of β-Thalassemia** |  |  |
| Genotype | $\beta^0/\beta^+$ | $\beta^E/\beta^+$ |
| Mutation | IVS-II-654*/ CD17 | CD26/CD41-42 |
| Average transfusion volume of 1 year prior to reinfusion (ml/kg/time) | 14.3 | 12.3 |
| Transfusion interval (Days) | 20 | 20 |
| Baseline of Hb (g/dL) | 12.8 | 9.2 |
| **Iron metabolism** |  |  |
| Ferritin (ng/ml) | 1486.79 | 1695.98 |
| Liver T2* MRI (ms) | 11.61 | 14.52 |
| Cardiac T2* MRI (ms) | 20.91 | 30.09 |
| Iron chelation | Deferasirox<br>800mg/day | Deferasirox<br>1000mg/day |

\* IVS-II-654 has been considered as $\beta^0$ mutation as well

### CD34^+^ cell harvest and transduction

In the first mobilization cycle, F001 and F002 underwent HSC mobilization with combination of G-CSF and Plerixafor, and G-CSF alone to explore preferable mobilization regimen. After the first round of mobilization and apheresis, insufficient amount of CD34^+^ cells were harvested from F002, so F002 experienced the second mobilization cycle with G-CSF and Plerixafor. The total number of 4.1 × 10^8^ and 3.5× 10^8^ CD34^+^ cells was collected from F001 and F002 patients (Table 2). In the meanwhile, the adequate backup CD34^+^ cells were cryopreserved for each patient according to the protocol requirement. The vector copy number (VCN) in the HGI- 001 injection of F001 and F002 was 3.36 and 3.10 respectively. The percentage of CD34^+^ cells in HGI-001 injection of F001 and F002 was 94.5% and 98.2%, and cell viability was 92.4% and 88.9% (Table 2).

**Table 2.**
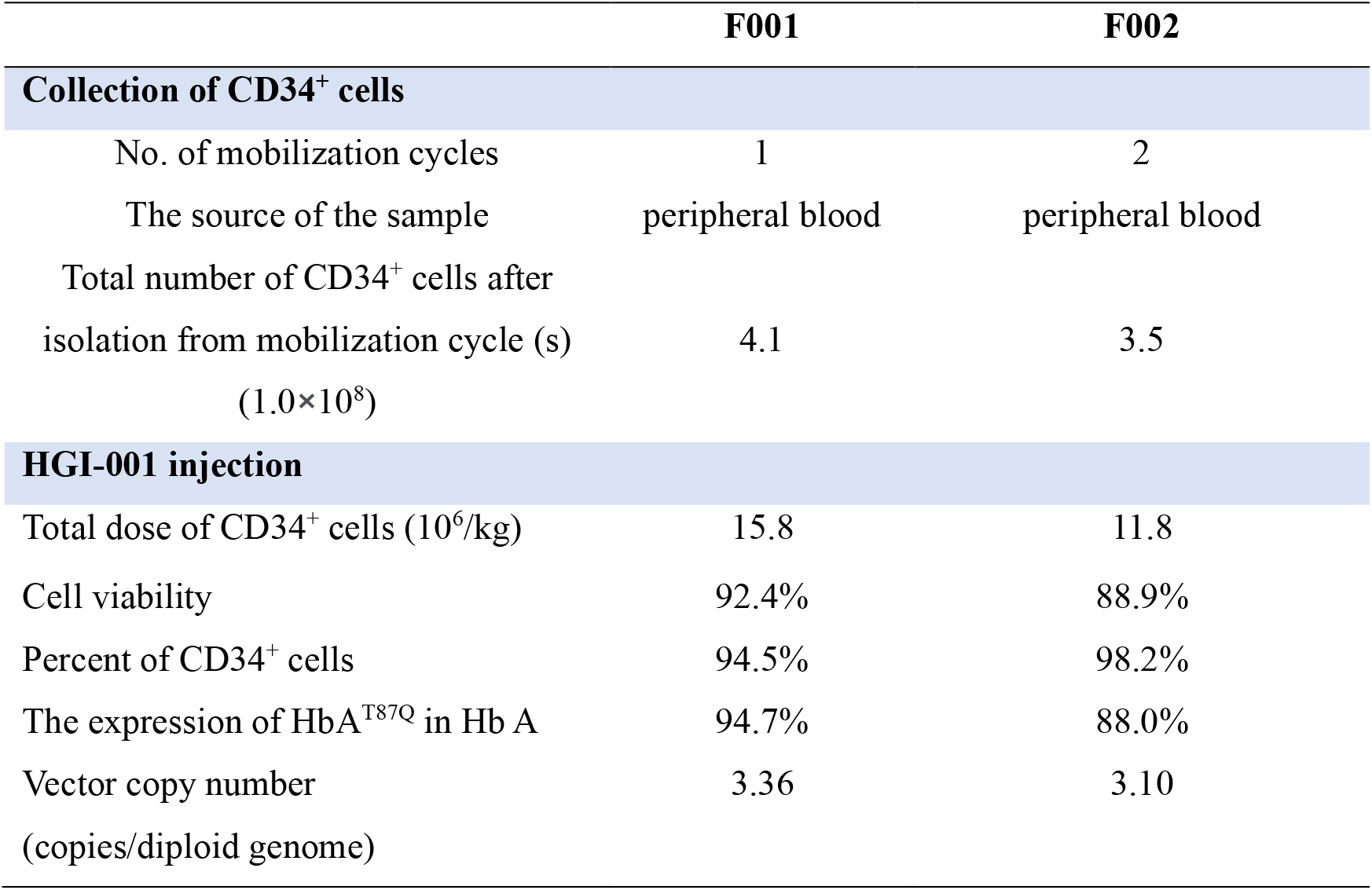
Collection of CD34^+^ cells and dosage of HGI-001 injection

### Conditioning and successful engraftment

Prior to the beginning of busulfan conditioning, iron chelation agents had stopped for at least a week to avoid the increased busulfan blood concentration by Deferasirox^24^. Patients’ conditioning was initiated with busulfan via intravenous infusion at 0.8mg/kg every 6 hours. The busulfan dose was adjusted depending on the first dose pharmacokinetics to reach expected targeted concentration. The area under the curve (AUC) of the first dose busulfan of two patients were 943.97 μM*Min and 1982.89 μM*Min. Patients received respective HGI-001 injection with the total dose of 15.8 and 11.8 million CD34^+^ cells per kg (Table 2).

The criteria for successful engraftment in the present study were that the absolute neutrophil count (ANC) was maintained above 0.5×10 ^9^/L for three consecutive days and the platelet count (PLT) was maintained above 20×10 ^9^/L for seven consecutive days without the support. After HGI-001 reinfusion, the time of neutrophils engraftment taken in patients F001 and F002 was 17days and 19 days; and the platelet engraftment spent 15 days and 13 days respectively (Figure 1A). The neutrophil and platelet number of F001 restored to the normal range at the discharge from transplant unit and the first month after reinfusion (Figure 1B). The ANC and PLT for F002 patient were still less than the reference range for Chinese children of corresponding age when discharged from transplantation unit, but increased significantly from one month after reinfusion and returned to the corresponding reference range from the sixth month after reinfusion (Figure 1B).

**Figure 1.**
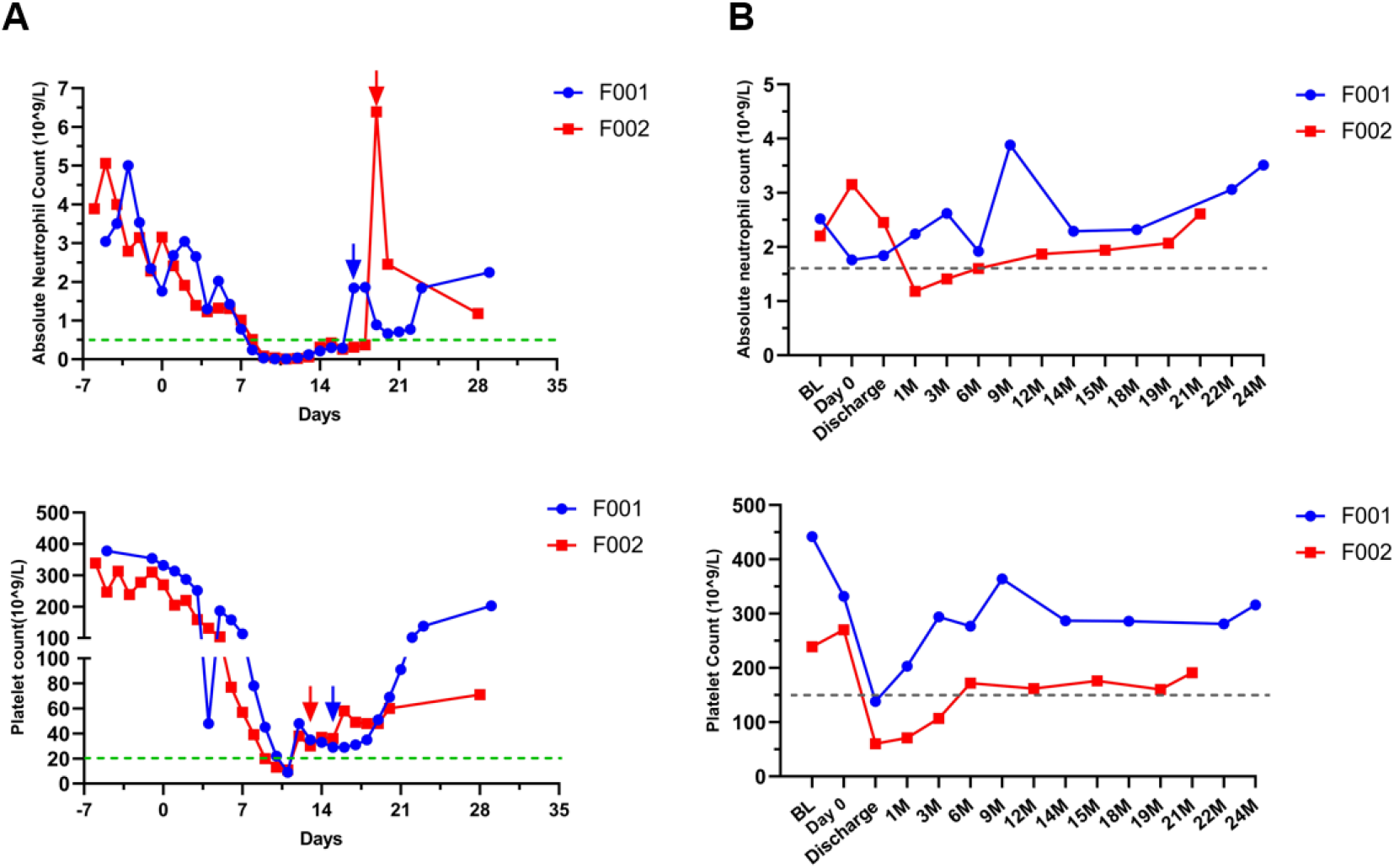
The successful bone marrow engraftment and the normalization of neutrophil and platelet counts occur in F001 and F002 patients. (A)Shown are alterations of neutrophil and platelet number from busulfan conditioning to successful engraftment after HGI-001reinfusion in both patients. (B) After successful engraftment, ANC and PLT have maintained the reference values until the most recent visit. The green and grey dashed horizontal lines respectively indicate the standards of engraftment in Figure 1A and the reference values in Figure 1B. Day 0, the day of HGI-001 reinfusion; BL, baseline.

### The efficacy of HGI-001 injection Transfusion independence

Patients F001 and F002 have completed follow-up visits in the 24 months and 21 months after gene therapy. Both subjects remained free from red-cell transfusions as of their most recent visits after discharge from the transplant unit. The last transfusion of F001 and F002 both occurred on the 13^th^ day coincidentally after reinfusion of drug product. Their average hemoglobin levels of F001 and F002 between 1 month after treatment and the last visits were 9.9 and 10.4 gram per deciliter respectively, both have achieved the transfusion-independent status as defined by this study. Furthermore, their survival time of independent transfusion was more than 720 and 630 days individually (Figure 2).

**Figure 2.**
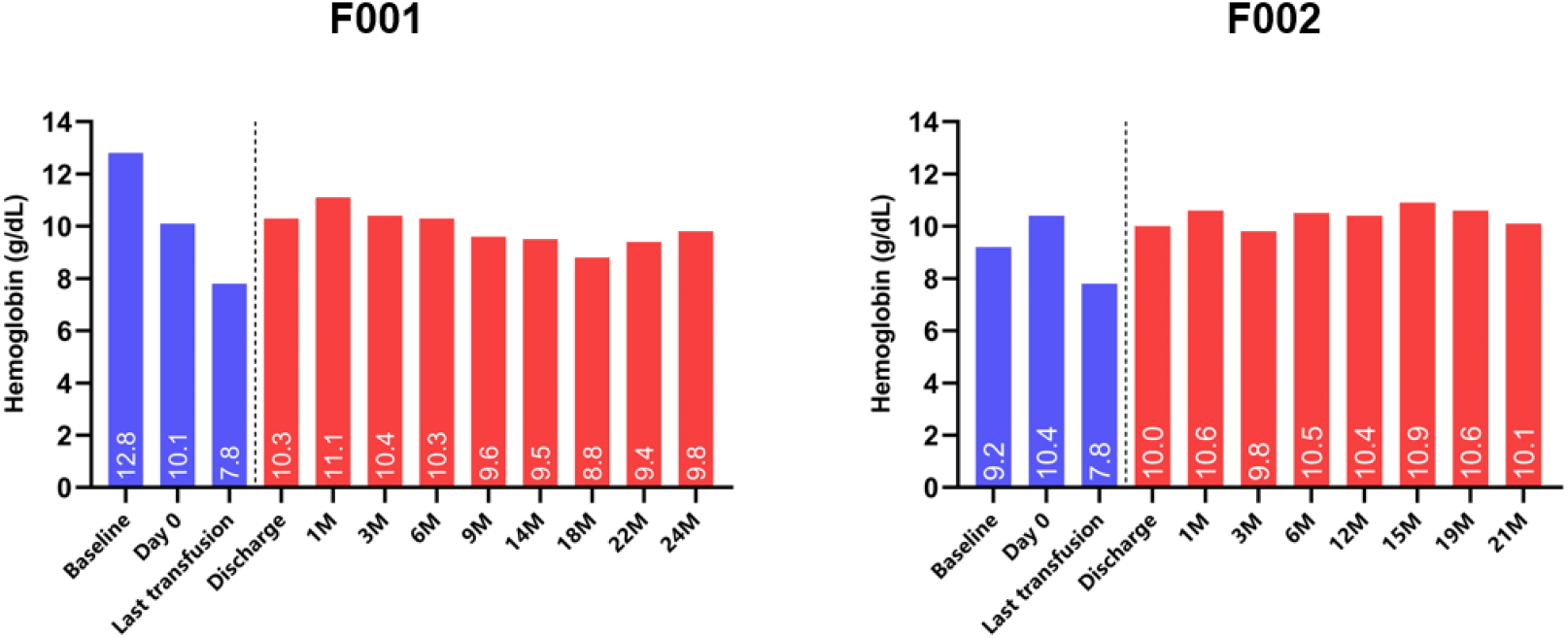
The total hemoglobin level in patients after HGI-001 injection reinfusion. Transfusion independence have occurred in both patients. The total Hb levels of F001 (left panel) and F002 (right panel) are displayed from screening (baseline) to the last follow-up visits. Total Hb levels at each follow-up visit are displayed in white letterings respectively. The blue and red bars represent the Hb level during the period when blood transfusion is needed or stopped.

### The kinetics of HbA^T87Q^ and VCN levels after gene therapy

The average levels of HbA^T87Q^ expression in F001 and F002 patients were 7.1 g/dL (range:6.6-7.5g/dL) and 7.0g/dL (range:6.0-7.3g/dL) between 3 months post-infusion and the last visit (Figure 3A). Until the last follow-up visit, the respective median VCN of F001 and F002 in PBMCs were maintained at the average of 1.22 (range: 0.97-1.88) and 2.11 (range: 1.21-2.42, Figure 3B). Additionally, efficient LentiHBB^T87Q^ vector expression could be persistently observed in F001 and F002 bone marrow samples (Figure 3B). As shown in Figure S5, the level of HbA^T87Q^ in both participants emerged remarkable ascending trends in the first three months after gene therapy because of the initiation expression of transduced exogenous gene (called ascending phase). Following the six-month gene therapy, the levels of HbA^T87Q^ expression in F001 and F002 were relatively unchanged (named plateau phase). These data have indicated that the kinetics of HbA^T87Q^ can generally increase throughout the 6 months after which time they stabilize.

**Figure 3.**
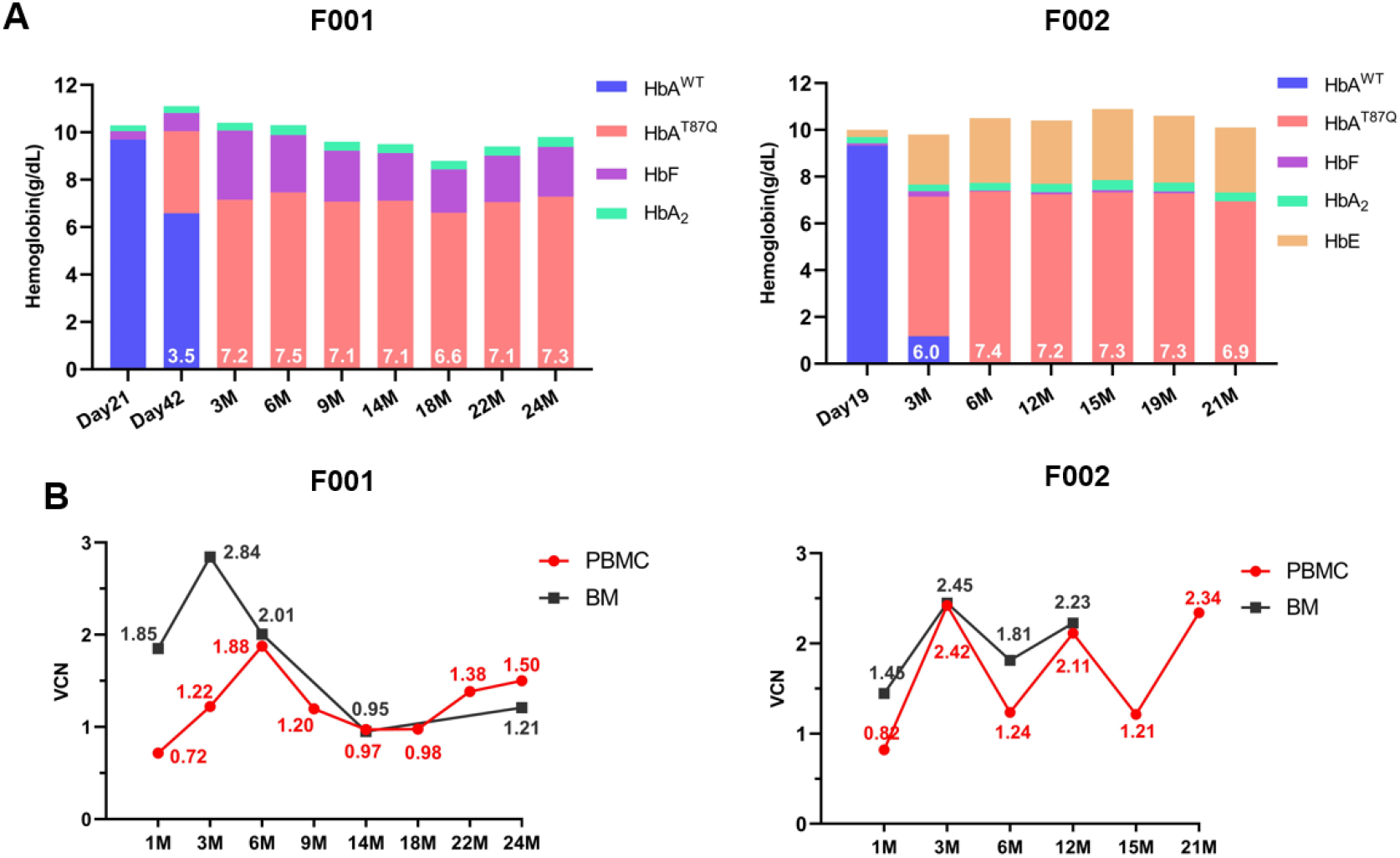
The kinetics of different hemoglobin types, and VCN of PBMC and BM in patients. (A) Hb concentrations (derived from β-like globin chain quantification) in patient’s peripheral blood, as assessed by HPLC analysis. Day, the days after HGI-001 reinfusion; M, planned protocol visits in months after reinfusion. HbA^T87Q^ concentrations are shown in white lettering. (B) Vector copy number (VCN) per copies genome in PBMC and marrow bone (BM) after reinfusion. The VCN values are shown in red (PBMC) and black (BM) lettering.

It is interesting to observe that in these two patients who have been transfusion independent, Hb F of patient F001 and Hb E of patient F002 also contributed to the total hemoglobin. The Hb F level of patient F001 reached a peak (2.9 g/dL) at 3 months after the reinfusion, then exhibiting a gradual downward variation sequentially (Figure S5). The genetic test revealed mutations at *BCL11A* and *HBS1L* genes in patient F001, indicating that they might be associated with an elevated level of Hb F (Table S2). As the patient F002 carried a β^E^ allele, endogenous Hb E level varied in a similar manner to HbA^T87Q^, with a sharp upward trend until the sixth month after gene therapy and a steady trend thereafter (Figure S5).

### The iron overload was relieved after reinfusion of HGI-001 injection

Patients are expected to eventually relieve iron overload when subjects express sufficient HbA^T87Q^ to achieve transfusion-free condition. Compared to pre-treatment, both subjects showed a significant reduction in iron level following gene therapy. The baselines of iron levels were 47.7μmol/L and 38.7μmol/L for F001 and F002. Iron levels were back to normality in both subjects from 3 months post-treatment, with F001 and F002 being 18.2μmol/L and 16.6μmol/L at the latest visit (Figure 4A). Serum ferritin levels for F001 and F002 were 798.0 ng/mL and 1016 ng/mL respectively at the Month 24 and Month 21 after HGI-001 reinfusion. Although they remained above the age-specific reference range, ferritin levels for F001 and F002 were considerably reduced over time compared to the baselines. There were transient peaks in serum ferritin at the first month after reinfusion, which have been speculated to be related to the continuously supportive red-cell transfusions in the transplant unit. After the patients were freed from blood transfusion, serum ferritin levels dropped sharply without iron chelation (Figure 4B). In addition, the MRI result of F001 at 24 months post-treatment revealed no abnormality in iron content in heart and liver tissue (Cardiac T2* MRI=26.84ms, Liver T2* MRI=7.15ms).

**Figure 4.**
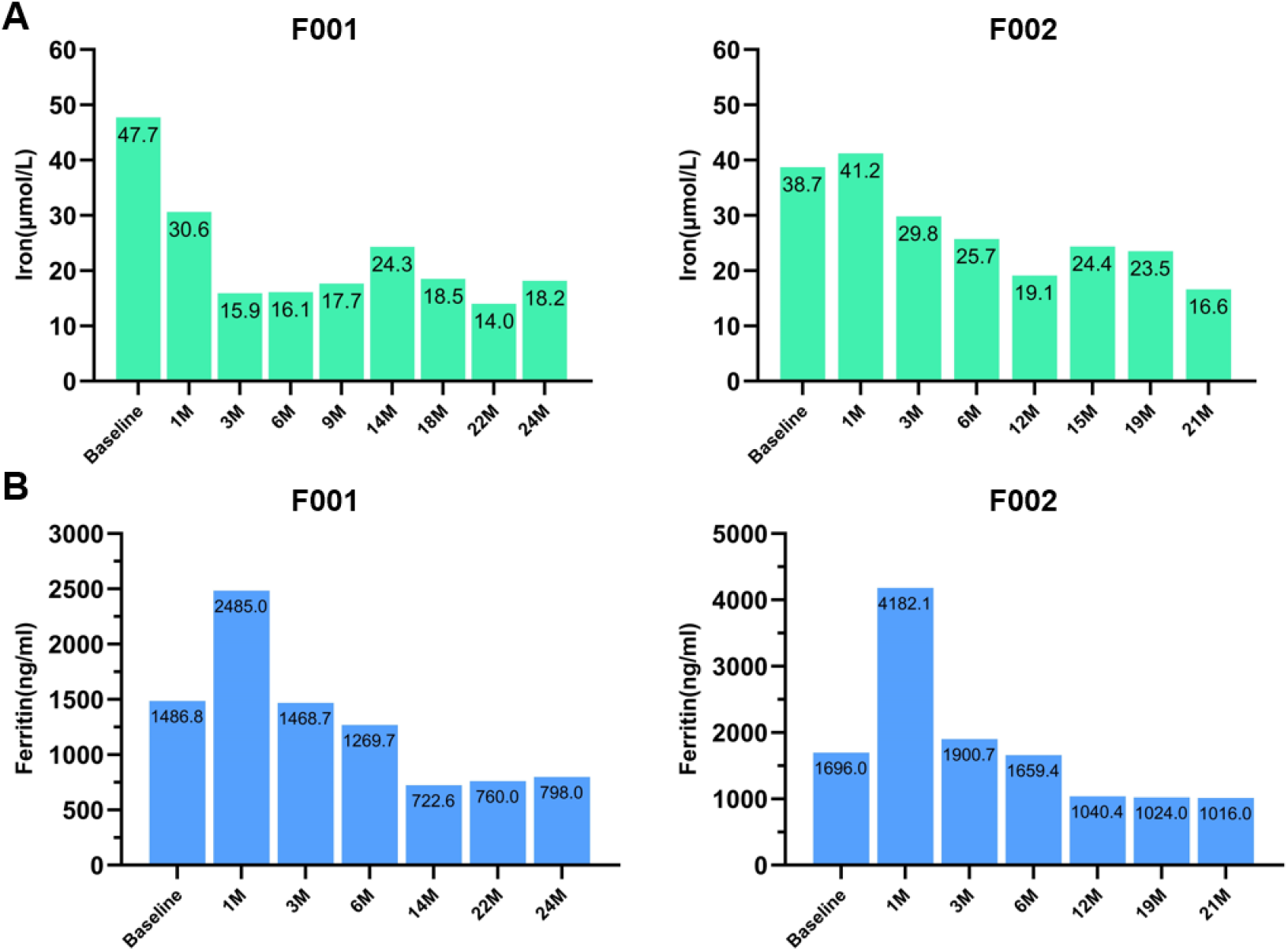
The transient reduction of iron burden is shown in patients. (A) The alterations of iron before and after reinfusion when patients became transfusion independent and discontinued iron chelation. (B) The alterations of serum ferritin before and after reinfusion. The numbers in black within the green and blue bars indicate the concrete values of iron and serum ferritin respectively.

### The changes of erythropoiesis and hemolysis

The levels of soluble transferrin receptor (sTfR) and erythropoietin (EPO) in serum are already known as the markers of ineffective erythropoiesis^25^. We investigated biomarkers of ineffective erythropoiesis after gene therapy. For F002 patient, the levels of sTfR and EPO were within normal range after stopping blood transfusion (Figure 5A). These results suggest that abnormalities in erythropoiesis were abolished in F002 with the β^E^/β^+^ genotype. In contrast, complete normalization of biomarkers of ineffective erythropoiesis was not observed in patient F001 (IVS-II- 654*/CD17) (Figure S7). The values of non-conjugated bilirubin declined to the normal reference range, indicating that hemolysis status of F001 and F002 was relieved after gene therapy (Figure 5B). Lactate dehydrogenase, which is a clinical marker linked to intravascular hemolysis, remains the normalization pre- and post- reinfusion of HGI-001 injection.

**Figure 5.**
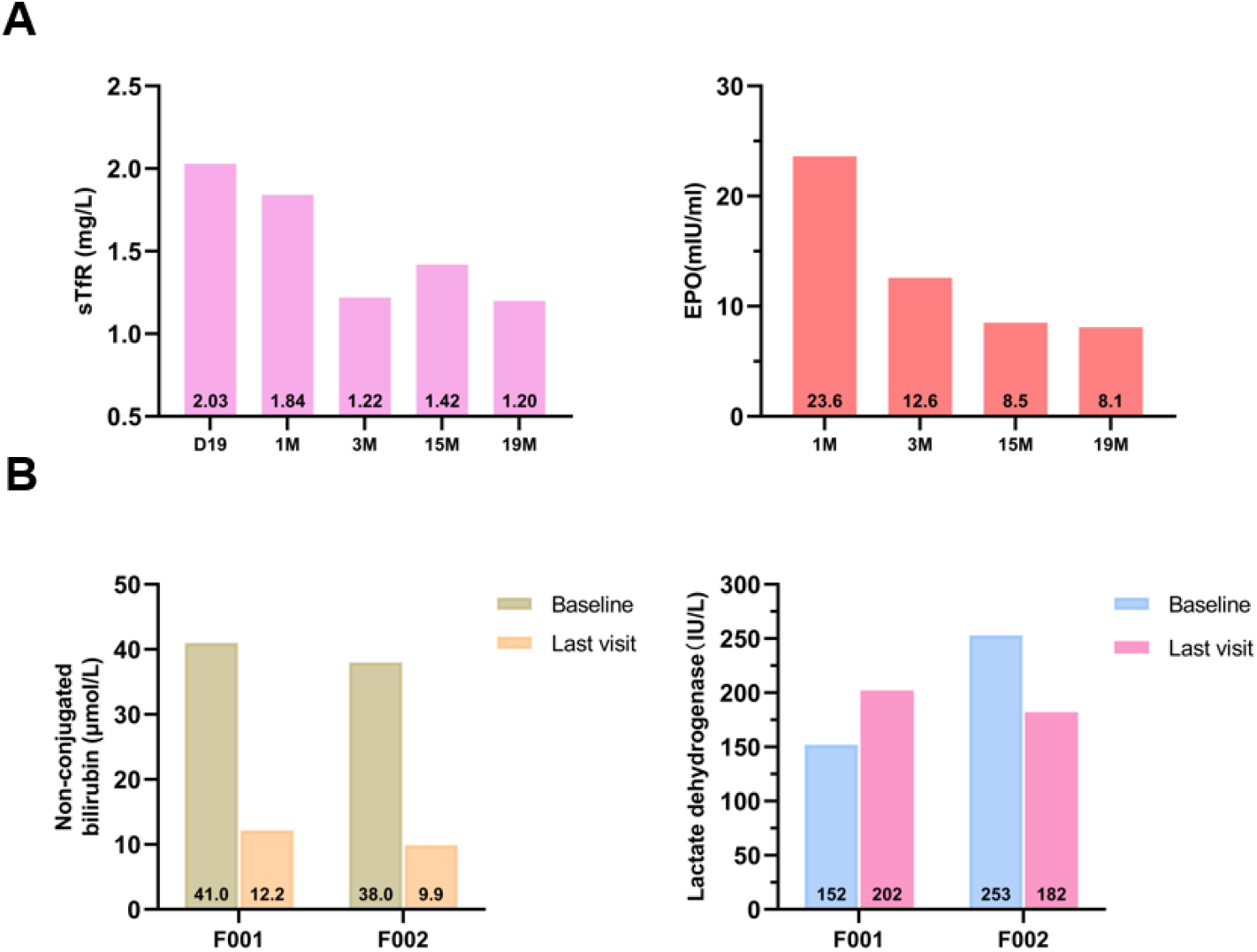
The alterations of soluble transferrin receptor (sTfR) and erythropoietin (EPO) levels in patients after HGI-001 injection reinfusion. (A) The levels of sTfR (left) and EPO (right) are normalized in F002 patients after HGI-001 injection treatment. (B) Non-conjugated bilirubin (left) and lactate dehydrogenase (right) of F001 and F002 patients return to normality. The reference range used here is sTfR (0.8-1.8mg/L), EPO (1.5-31.9mIU/ml), non-conjugated bilirubin (2.0-17.0μmol/L) and lactate dehydrogenase (157-272IU/L) respectively. The numbers in black within the bars indicate the concrete values.

### Safety assessment

#### No clonal dominance is discovered through integration site analysis

In normal, especially post-transplantation hematopoiesis, clonal dominance was thought to be an uncommon but potential occurrence and an important outcome of malignant progression^26^. The vector-related adverse events include insertional mutagenesis and replication competent lentiviruses (RCL)^27^. Integration site analysis (ISA), which has been a necessary bioinformatic method recommended by FDA, was applied to monitor the insertional oncogenesis. The number of vector insertion sites (VIS) was 5017 and 1539 in PBMC sample respectively from F001 at 21 months and 15 months after HGI-001 injection reinfusion. The proportion and annotation of the top 10 insertion sites are demonstrated in Figure 6A. In the peripheral blood samples, the distribution of ISA in the genome and the preference of upstream and downstream bases are in line with the distribution of insertion sites of lentiviral vectors (Figure 6A). We have not found clonal dominance through ISA analysis in our study. Moreover, no abnormal blood cell proliferation was found in the bone marrow of F001 and F002 after gene therapy (Figure 6B and Figure S6).

**Figure 6.**
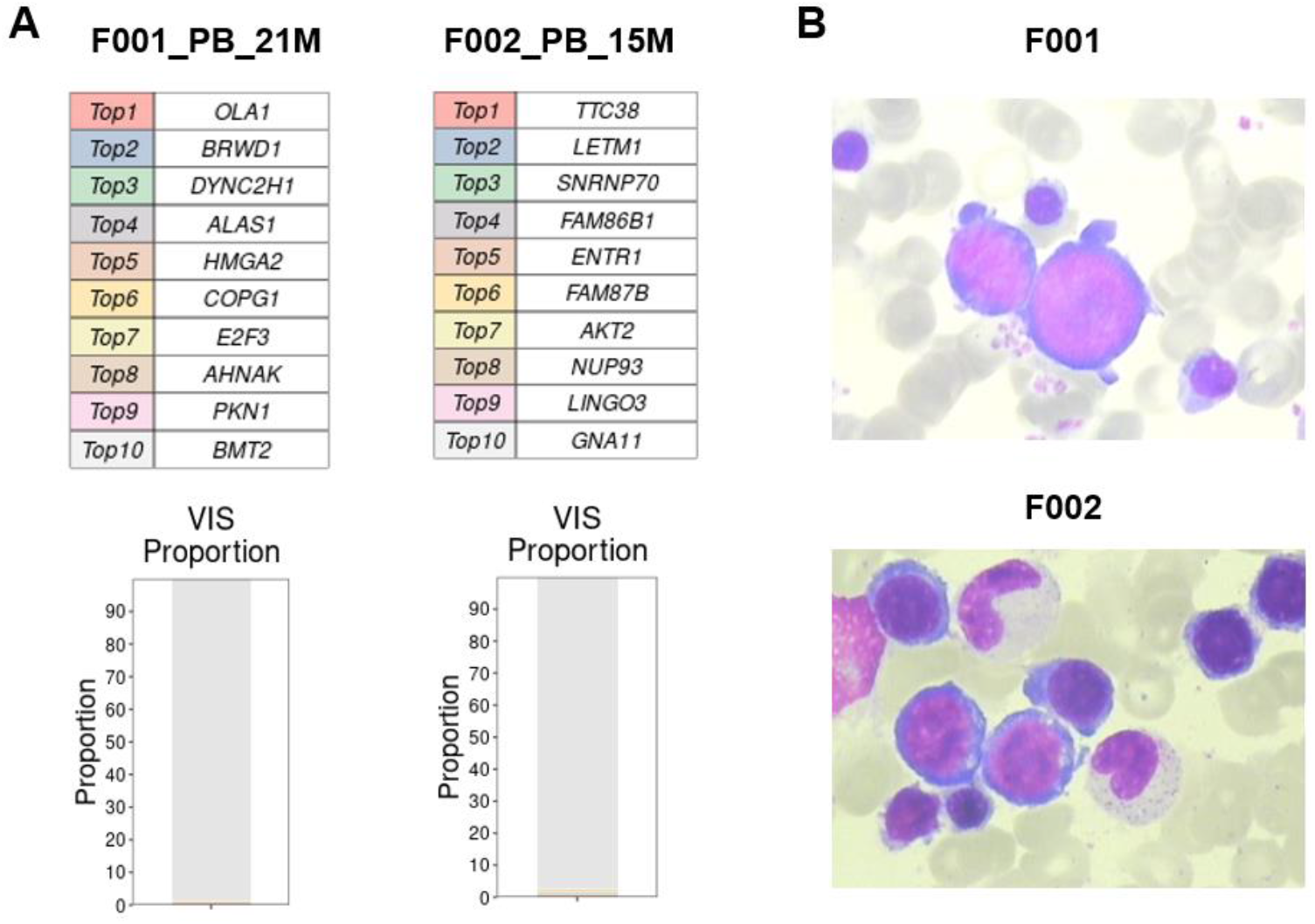
No clonal dominance has been discovered via Integration Site Analysis (ISA) and bone marrow smear for the patients. (A) Top 10 insertion sites are shown by ISA. (B) The upper panel is shown the image of marrow swear of F001 in the 14 months after reinfusion; the lower panel is the image of marrow swear of F002 in the 12 months after reinfusion.

#### Other adverse events of F001 and F002 in this study

No serious adverse event (SAE)/deaths/other adverse events of special interest are observed till now. There were 27 AEs in subjects F001 and F002 between the signing of the informed consent form and the most recent visit, including 4 AEs in the period of myeloablative conditioning, 21 AEs in the period of transplant unit, and 2 AEs after discharge (Table S3). The most AEs occurred during the transplant unit phase, accounting for 77.8% (21/27), and the AEs of severity ≥Grade 3 accounted for 47.6% (10/21), like febrile neutropenia, neutropenia, leukopenia, thrombocytopenia, pharyngalgia, and oral thrush. The only AE considered probably drug-related was engraftment syndrome (Grade 1). Moreover, hepatic veno-occlusive disease (VOD) limited to Grade 1 occurred to patient F001 on Day 12 and Day 48 after reinfusion of HGI-001 injection, which has been well known as a side effect caused by busulfan conditioning. All adverse events have recovered after clinically standard therapeutic management.

#### The lymphocyte reconstitution after gene therapy

The CD3^+^, CD4^+^, CD8^+^ T lymphocyte, B lymphocyte of patient F001 and F002 were reconstituted rapidly, which did not differ obviously from normal reference range at discharge. However, the reconstitution of natural killer (NK) cells was delayed compared with other lymphocyte subsets, and the counts and proportion of NK cells were still lower than normal range when the patients discharged from the transplant unit (Figure 7). At the first and third month after gene therapy, the number of NK cells in F001 and F002 was upregulated to 189/μL (12.24%) and 103/μL (9.98%), which the normalization of NK cells respectively occurred.

**Figure 7.**
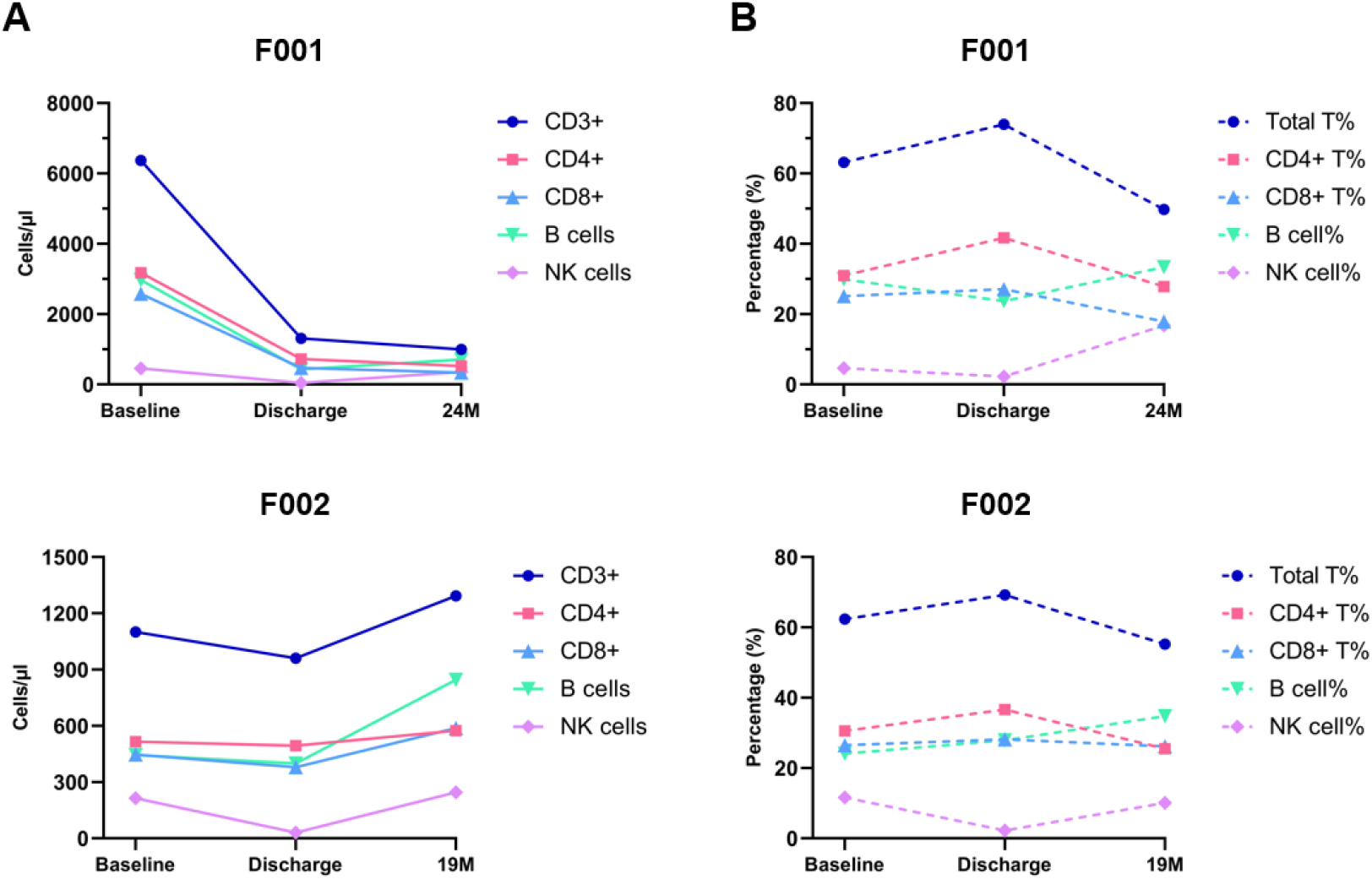
The lymphocyte subset reconstitution in the peripheral blood for F001 and F002 patients after HGI-001 reinfusion. (A) The number of lymphocyte subsets and (B) the proportion of lymphocyte subsets in patients F001 and F002 between screening and the last follow-up visit. Baseline refers to the data of lymphocyte subpopulation test at the screening phase. Discharge is defined as patients leaving the transplant unit after successful engraftment and clinical stability.

## Discussion

We demonstrate in this study the safety and efficacy of gene therapy for TDT using the clinical grade LentiHBB^T87Q^ (cLentiHBB^T87Q^) vector with a follow-up period of 24 and 21 months respectively. Currently, F001 has completed all of follow-up visits in this trial and achieving the primary efficacy endpoint. Both of F001 and F002 patients have achieved the secondary effective endpoint of this study, having not been transfused for at least twelve consecutive months with a mean Hb ≥ 9.0 g/dL after HGI-001 injection treatment.

HGI-001 injection was administrated after complete myeloablation conditioning in two pediatric patients with TDT. The busulfan was selected to administer for myeloablation conditioning referred to the other ongoing or completed gene therapy clinical study ^18,28^. A high busulfan exposure is strongly correlated with an increased risk of hepatic toxicity and non-relapse mortality. VOD, which is a busulfan-related clinical problem, is paid the special attention when busulfan-based conditioning HCT^29^. According to the institutional standards, ursodesoxycholic acid and heparin sodium are administrated starting at the initiation of busulfan conditioning and continuing at least 3 weeks. Patient F001 developed VOD twice limited to Grade 1, and VOD was ended soon after standard treatments in clinic. The occurrence of VOD appears to be incompletely related to the cumulative level of busulfan exposures. The AUC of the first dose busulfan of F002 was much higher than that of F001 and our recommendation, as well more than the current target exposures in the EMA and FDA labels of busulfan. In another clinical trial of gene therapy patients with TDT, VOD appeared in four patients (grade 4 or grade 2) after busulfan conditioning^18^, while patients with liver VOD had approximated average daily busulfan AUC values that were comparable to patients without VOD. Nevertheless, we still recommend the close monitoring the busulfan exposures during myeloablation conditioning. Unless patients have contraindications, all patients should receive VOD prophylaxis before the beginning of busulfan conditioning.

No SAEs, AEs of special concern for lentiviral gene therapy or unexpected complications were observed in relation to HGI-001 injection. We regularly monitored the vector integration sites by ISA and replication-competent lentivirus by detecting the gene of VSV-G protein via PCR assay. To date, no evidence of vector-related clonal dominance or cancer cases have been found in this study, nor has RCL. The risk of the occurrence of insertional oncogenesis was identified in gene therapy studies with γ-retroviral vectors^30,31^. Hence, lentiviral vectors have turned out to be the current preference for most hematopoietic stem cell gene therapy^18,20,28,32^.

The only drug-related AE is engraftment syndrome (ES) limited to grade 1, which started on Day 11 and ended on Day 13 after reinfusion. ES is a clinical complication during neutrophil recovery occurring more frequently following auto-HSCT. The administration of corticosteroid treatment is also effective for ES in our trial. The duration of platelet engraftment is particularly critical for patients with TDT, as platelet transfusion are often ineffective with high-risk for bleeding while in conditioning-related aplasia^33^. Delayed platelet engraftment is observed in the Zynteglo treatment (a phase 3 study, NCT02906202), which is a similar drug product to HGI-001 injection, and the median of successful platelet engraftment is 46 days (from 20 to 94 days) after Zynteglo administration^18^. In our study, the platelet could be engrafted successfully on Day 15 and Day 13 for F001 and F002 patients respectively. In addition, three new subjects were treated with the upgraded HGI-001 injection, the results of which are shown in Figure S4. With these five patients all together, the median of successful platelet engraftment was 15 days (range:13 to 20 days). During nearly 24-month follow-up visits, both patients remain remarkably medically stable. An upper respiratory infection (URI) occurred to F001 accompanied with fever (39.5°C) and sight cough on Day 679. Patient F001 did not receive treatments and spontaneously recovered three days later. It is indicated that the lymphocyte subpopulations and function can reconstitute successfully after HGI-001 injection refusion.

The goal for TDT patients who received HGI-001 injection is to allow for durable transfusion independence, which we have achieved in both patients. Patients F001 and F002 have discontinued receiving the blood transfusion for up to 24 and 21 months, maintaining the weighted average Hb level of 9.9g/dL and 10.4g/dL respectively. It is well known that LentiHBB^T87Q^ gene therapy for transfusion-dependent β-thalassemia patients requires erythroid-specific and sustained expression of exogenous HbA^T87Q^ in erythrocytes^22^. The current data demonstrate that HGI-001 injection produces potent lentiviral gene vector-derived HbA^T87Q^. Approximately three months after HGI-001 injection reinfusion, HbA^T87Q^ derived from drug product was rapidly elevated, gradually replacing hemoglobin obtained by patients from blood transfusions. After 6 months of treatment, HbA^T87Q^ levels became relatively stable and have continued until the last visit, suggesting the clinical efficacy and stability of HGI-001 injection. In our clinical trial, we prove that cLentiHBB^T87Q^ that we previously established enables β-globin expression in HSPCs-derived erythrocytes from TDT patients^22^.

According to previous studies, the lowest related transfusion-independent VCN values is 0.8 in peripheral blood and bone marrow samples with GLOBE vector^19^. In addition, 7 of 22 cases expressed HbA^T87Q^ >8g/dL and they received VCN>0.6 drug product and all these cases have arrived the endpoint of transfusion-independent status in HGB-204 and HGB-205 trials^28^. We are currently unable to demonstrate an association between VCN in drug products and clinical benefit, but persistently high levels of VCN in peripheral blood and bone marrow cells appear to be a great contributor to drug-product efficacy regardless of the genotype of β-thalassemia.

Hb F in patient F001 also contributes to their total hemoglobin. The sequence of the functional factor regulating the level of Hb F or the number of F cells (Hb F-containing erythrocytes) is in or very close to the 2nd intron sequence of the 3kb *BCL11A* gene between the rs1427407 and rs4671393^34^. Sequencing reveals that patient F001 has mutations in the *BCL11A* gene at the rsl427407, rs766432 and rs4671393. A correlation between the rs4671393 of the *BCL11A* gene and >10% Hb F expression in pediatric patients has also been reported^35^. This evidence explains the sharply climbing percentage of Hb F in patient F001 after 3 months of treatment, followed by a slight decrease and then a stable trend. Elevated level of Hb F caused by point mutations in rs1427407, rs766432, and rs4671393 in the *BCL11A* gene of patient F001 does not seem to affect the expression of HbA^T87Q^. Since patient F002 genetically carries the β^E^ allele, the kinetics of endogenous Hb E parallels that of HbA^T87Q^ reaching plateau levels at the 6 months after HGI-001 injection reinfusion.

The reduction of iron load is considered as a slow and continuous process. In our study, these two subjects stopped taking iron chelated drugs one week prior to busulfan conditioning. The long-term absence of blood transfusion has contributed to alleviation of iron overload in both patients who were treated, showing the normalization of iron level and an obvious declining serum ferritin. The cardiac and hepatic T2* for F001 by MRI are normal at screening enrollment and the 24 months after gene treatment. As iron accumulation is difficult to reverse, patients with severe iron overload should be excluded from the ongoing gene therapy clinical trials for safety reasons^36^. Dys-erythropoiesis and hemolysis, the two pathological features of thalassemia, have been both eliminated in F002 patients with β^E^/β^+^. In contrast, the abnormities of clinical hallmarks of dys-erythropoiesis have persisted in patient F001 (IVS-II-654/CD17), whereas the correction of hemolysis can be observed. The similar cases reported in BB305 vector trial, ineffective erythropoiesis of 3 patients with β^E^/β^0^ was reversed, the patient with IVS1-110 homozygote still showed the dyserythropoiesis evidence^28^.

Here, we report only efficacy and safety data of F001 and F002. The follow-up of three additional subjects who have received the upgraded HGI-001 injection is ongoing (Table S4). The additional three subjects have been transfusion-free for more than 9, 6, and 2 months, respectively. The upgraded HGI-001 injection appeared to result in higher HbA^T87Q^ expression and higher total Hb in these three patients compared to subjects F001 and F002. In summary, HGI-001 injection can treat transfusion-dependent β-thalassemia and achieve transfusion independence. The long-term efficacy and safety of HGI-001 injection, as well as the differences of upgraded technologies remain to be seen.

## Data Availability

All data produced in the present work are contained in the manuscript.

## Contributors

<Sixi Liu and Chao Liu> conceived and designed the clinical trial. <Yue Li > executed the research, collected, and interpret data. <Nan Han> contributed to the data analysis, and writing and editing of the manuscript. <Wenjie Ouyang> contributed to the vector design. <Guoyi Dong, Xinru Zeng and Huilin Zou> contributed to manufacturing HGI-001 injection. <Honglian Guo> contributed to the quality control and releasing of HGI-001 injection. <Yan Huang and Nan Han> contributed to patient follow-up and data collection. <Yue Chen> contributed to the ISA of bioinformation and data analysis.<Wenwen Yao and Jiajun He> contributed to the detection of VCN and HbA^T87Q^. <Sixi Liu and Chao Liu> contributed revision of the manuscript. All authors had full access to raw data and had final responsibility for the decision to submit for publication. All authors reviewed, and approved the final version of this manuscript.

## Acknowledgements

We sincerely thank the support provided by China National GeneBank.

## Ethics approval and consent to participate

Permission for this study was obtained from the Medical Ethics Review Committee of Shenzhen Children’s Hospital (NO.202000210), and the Institutional Review Board of BGI (NO. BGI-IRB 20155-T3).

