## Supplementary Appendix for "Gene therapy using optimized LentiHBB^T87Q^ vector in two patients with transfusion dependent β-thalassemia"

### 1. The overview of HGI-001 injection clinical trial protocol

There are seven main steps in the HGI-001 injection treatment in this trial. The treatment process includes screening, mobilization and apheresis, HGI-001 injection manufacturing, myeloablative conditioning, reinfusion of HGI-001 injection and 24-month follow-up visits. Each subject will require approximately 30 months from the signing of the informed consent form to the completion of the last visit.

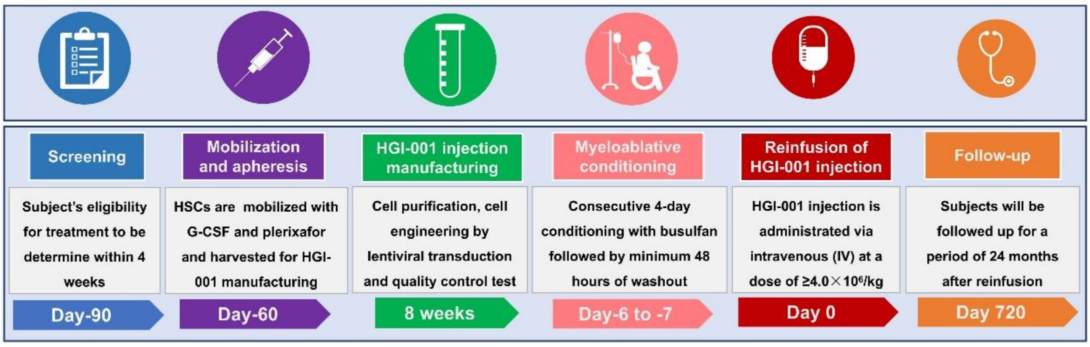


**Figure S1. The main steps in the process of HGI-001 injection clinical trial protocol.**

### 2. Inclusion and Exclusion criteria

Table S1 The inclusion and exclusion criteria in this trial

| | |
|---|---|
| **Inclusion Criteria:** | **1.** Participants between 8 and 16 years of age at the time of consent or assent (as applicable). For patients under the age of 18 years, written informed consent is provided by a parent or guardian. |
| | **2.** Diagnosis of transfusion dependent beta-thalassemia (TDT) without restriction of genotypes of subjects. |
| | **3.** At least 100 mL/kg/year of packed red blood cells (pRBCs) or ≥8 transfusions of pRBCs in the 2 years preceding enrollment (all subjects). |
| | **4.** Adequate transfusion for at least 3 months prior to screening to maintain hemoglobin (Hb) level ≥9.0 g/dL. |
| | **5.** The level of serum ferritin <3000μg/L; Cardiac magnetic resonance imaging (MRI) T2* >10ms and Liver MRI T2* >1.4ms; (subjects can provide records of iron chelation therapy within 3 months prior to screening). |
| | **6.** Clinically stable, and eligible for autologous hematopoietic |

| | |
|---|---|
| | stem cell transplant (HSCT). |
| | **7.** Adequate organ function for the conditioning with busulfan. |
| | **8.** Treated and followed for at least the past 2 years in a specialized hospital (Subjects are eligible for follow-up, and complies with the clinical trial schedule of assessments). |
| **Exclusion Criteria:** | **1.** A known and available human leukocyte antigen (HLA)-matched family or unrelated donor |
| | **2.** Prior receipt of HSCT and gene therapy |
| | **3.** A history of splenectomy |
| | **4.** Uncorrected bleeding disorder |
| | **5.** Uncontrolled seizure disorder |
| | **6.** Presence of psychoactive substance, drug, or alcohol abuse within six months prior to screening. |
| | **7.** Previous use of Hydroxyurea, Ruxolitinib, Decitabine or Cytarabine within 3 months prior to screening. |
| | **8.** Pulmonary hypertension without effective intervention. |
| | **9.** Positive for blood cell antibody in antibody screening test. |
| | **10.** Positive for presence of human immunodeficiency virus type 1 (HIV-1) or 2 (HIV-2), hepatitis B virus (HBV), or hepatitis C virus (HCV), Cytomegalic Virus (HCMV) or Epstein-barr virus, and treponema pallidum antibody (TP-Ab).<br>**Note:**<br>**a.** Subjects with positive antibody are eligible due to they have been vaccinated.<br>**b.** Positive for hepatitis B surface antigen (HBsAg) and HBV DNA copy number > upper limit of normal value is excluded (HBV DNA copy testing is not required for those who are negative).<br>**c.** Where clinically and/or regionally indicated, other tests may be performed, in which case positive results would exclude the subject from participating: for example, human T-lymphotropic virus-1 (HTLV-1) or -2 (HTLV-2), tuberculosis, toxoplasmosis, etc. |
| | **11.** Any prior or current malignancy, myeloproliferative, immunodeficiency disorder or autoimmune disease. |
| | **12.** Immediate family member with a known or suspected Familial Cancer Syndrome (including but not limited to hereditary breast and ovarian cancer, hereditary non-polyposis |

| | |
|---|---|
| | colorectal cancer and familial adenomatous polyposis). |
| | **13.** Clinically significant and active bacterial, viral, fungal, or parasitic infection. |
| | **14.** Subjects with other medical conditions who are not eligible to participate in the study (such as advanced liver disease, kidney disease or cardiac disease, etc.).<br>**Advanced liver disease, defined as:**<br>**a.** Persistent aspartate transaminase, alanine transaminase, or direct bilirubin value >3× the upper limit of normal (ULN), or<br>**b.** MRI of the liver demonstrating clear evidence of cirrhosis, or<br>**c.** Liver biopsy suggests active hepatitis, significant fibrosis, conclusive evidence of cirrhosis (Liver biopsy is only carried out when MRI findings suggestive of active hepatitis, significant fibrosis, inconclusive evidence of cirrhosis).<br>**Severe kidney disease, defined as:** creatinine clearance rate< 30% of the normal value. |
| | **15.** A white blood cell (WBC) count $<3\times10^9$/L, and/or platelet count $<100\times10^9$/L. |
| | **16.** Diagnosis of significant psychiatric disorder of the subject that could seriously impede the ability to participate in the study. |
| | **17.** Subjects with diabetes, thyroid dysfunction, or other endocrine disorders. |
| | **18.** Participation in another clinical study with an investigational drug within 4 weeks of Screening. |
| | **19.** An assessment by the investigator that the subject would not comply with the study procedures outlined in the protocol. |
| | **20.** Any other condition that would render the subject ineligible as determined by the primary investigator, like a history of allergy to drug ingredients. |

**3. The protocol of mobilization and apheresis**

Hemoglobin level of subjects is recommended to be maintained ≥9.0 g/dL during the mobilization and apheresis phase. Human granulocyte colony-stimulating factor (G-CSF) injection is administrated with 5μg/kg every 12 hours (q12h) (Kexing Biopharm Co., Ltd., Specification: 75μg/ml/Syringe). Dosage of human G-CSF can be adjusted when the count of white blood cells is higher than $100\times10^9$/L before the day of apheresis. Plerixafor is administrated on the evening of Day 4 of mobilization, which is purchased from Sanofi-aventis. Apheresis is recommended on the morning of Day 5 of mobilization (the day after plerixafor administration). $CD34^+$ cell count is done by flow cytometry between the 3rd day and the 5th day (Figure S2). The specific beginning time is determined by the physicians. The appropriate physical examinations are carried out before the initiation and after completion of apheresis according to standard operation procedure of clinical site. There can include up three sequent apheresis days with in a mobilization cycle (from the $5^{th}$ to $7^{th}$ day of the mobilization cycle). The physician should follow the instructions of the peripheral mononuclear cell collector.

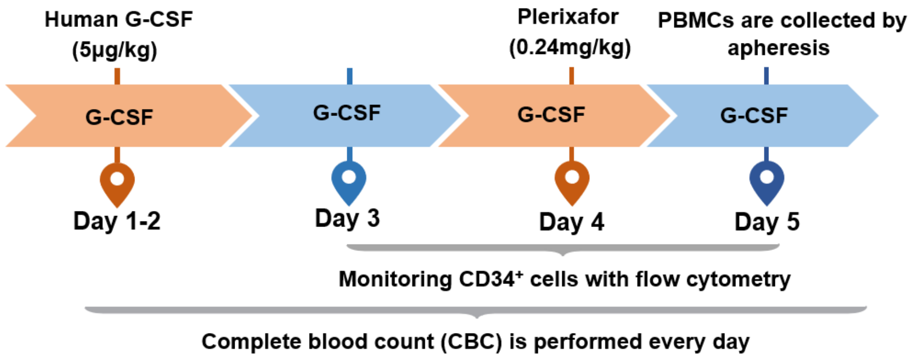


**Figure S2. The standard process of mobilization and apheresis.**

### 4. The standard protocol of myeloablative conditioning and reinfusion

The conditioning criteria include (1) HGI-001 injection to be used for a particular subject has been release tested, dispositioned for clinical use, and is stored at the clinical site; (2) the subject has received medical examinations and evaluation before conditioning; (3) iron chelation has been stopped at least 7 days prior to start of the conditioning.

As shown in Figure S3, the conditioning regimen in this study consists of four consecutive days of intravenous busulfan at a typical starting dose of 3.2 mg/kg/d. To reduce the risk of adverse events during myeloablative conditioning, it is recommended to administer the busulfan every 6 hours (q6h), divided into four doses of 0.8/mg/kg/day, with each time lasting approximately for two hours. Busulfan adjustments are made based on monitoring the pharmacokinetic on the first day and are determined by the physician as medical condition of patients. Additionally, CBC, serum chemistry and liver and kidney function tests need to be monitored daily throughout the myeloablative conditioning process in order to manage potential serious adverse events that may be caused by busulfan. Prophylaxis for epilepsy should be carried out 12 hours prior to conditioning. Additional prophylaxis may be performed depending on the subject's clinical condition, such as prevention of infections and hepatic veno-occlusive disease (VOD). The busulfan will be eliminated for a minimum of 48 hours to ensure that it is completely washed out. The need for additional treatment prior to HGI-001 Injection can be decided by the physicians depending on experiences and the subject's individual condition.

The HGI-001 injection is stored in gaseous liquid nitrogen. Before infusion, the package of HGI-001 injection should be checked for complete labeling, patient number, product information and appearance. Then, it should be thawed in 37 ± 1°C water bath for approximately one minute. The day of HGI-001 injection is defined as Day 0. If the subject undergoes more than one cycle of apheresis, there should be two or more packages of drug products, which should be administered sequentially.

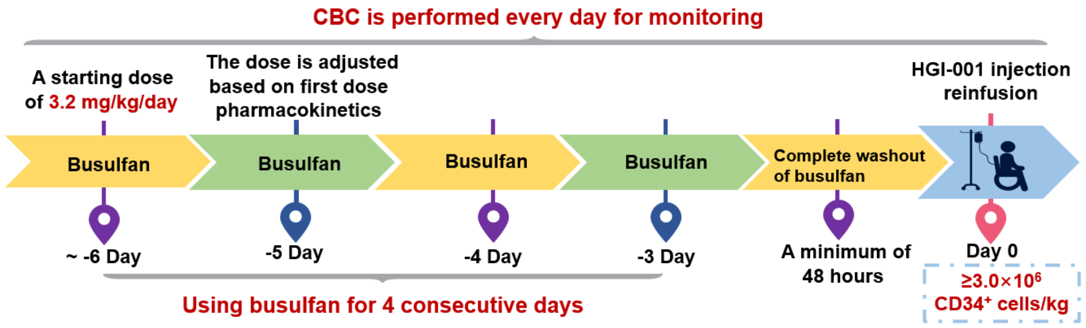


**Figure S3. The standard process of myeloablative conditioning and reinfusion. CBC: complete blood count.**

### 5. The method of Integration site analysis

Combining the LAM-PCR (linear amplification mediated PCR) and high-throughput sequencing techniques, genomic fragments containing the lentiviral integration sites were specifically enriched and sequenced as previously described. The genomic DNA extracted from patients' bone marrow of peripheral blood mononucleated cells were sonicated into 250-500bp fragments. Then adaptors containing a 10bp UMI each (unique molecular identifier) were ligated to the end-repaired DNA fragments. In order to identify and enrich the DNA fragments containing the integration sites, a LTR (long terminal repeat) specific and an adaptor specific primer were used to conduct a semi-nested PCR on adaptor ligated fragments. Then, another round of PCR was conducted to add 10 bp barcodes to both ends of the fragments to correctly label the DNA from different samples. The final PCR products from different samples were pooled in equimolar ratio and subjected to DNA nanoball generation. The constructed libraries were sequenced using the MGISEQ-20RS platform generating 150bp paired-end reads.

The sequenced reads were first filtered using SOAP nuke[1] to remove reads with low quality. Then custom scripts were used to trim the LTR segments and identify the UMI for each sequence. Then the pre-processed sequences were mapped to the vector genome to remove those from the lentivirus vector before mapping to the human genome (GRCh38.p12) using BLASTn (RRID:SCR_001598)[2] and HISAT2[3]. The integration sites were identified based on the alignment file and annotated using bedtools [4]. The relative abundance of the integration sites was calculated based on the sequence UMI counts.

### 6. Supplementary data

#### 6.1 The successful engraftment occurs rapidly after reinfusion with upgraded HGI-001 injection

The neutrophil and platelet engraftment of additional three subjects who were treated with upgraded HGI-001 injection. The neutrophil engraftment of patients S001/S002/S003 was occurred successfully on the Day 18, Day 22 and Day 16, and the platelet engraftment of these three patients was observed on the Day 18, Day 20 and Day 13 respectively (Figure S4).

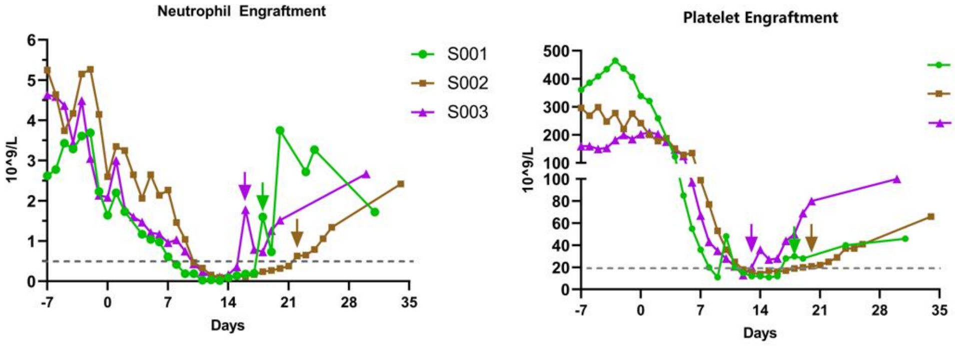


**Figure S4. The alterations of ANC and PLT in S001/S002/S003 patients with upgraded HGI-001 injection between the busulfan conditioning and successful neutrophil and platelet engraftment.** The grey dashed horizontal lines indicate the standards of engraftment.

### 6.2 Change in the proportion of each contributor in the total hemoglobin

Exogenous $HbA^{T87Q}$ increased rapidly in the first three months after reinfusion of HGI-001 injection, and hemoglobin adult from blood transfusion (called $HbA^{WT}$) decreased, which was obviously replaced by $HbA^{T87Q}$ in F001 and F002. From three to six months after the HGI-001 injection reinfusion, there was a small increase of $HbA^{T87Q}$ level, and then remained stable until the last follow-up visit .

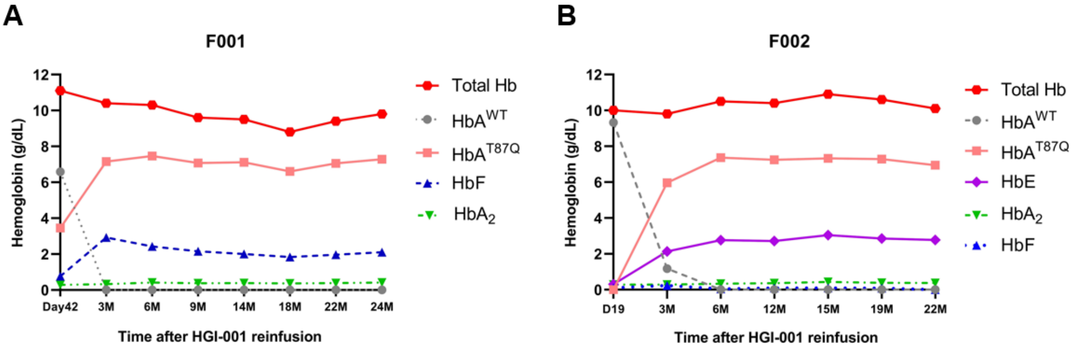


**Figure S5. The levels of each contributor in the total hemoglobin remain stable at 6 months after gene therapy.**

### 6.3 Table S2 List of F001 mutations in sites associated with Hb F regulation

| **Gene Symbol** | **Function** | **cHGVS** | **rs ID** |
|---|---|---|---|
| *BCL11A* | intron | c.386-17267T>C | rs10189857 |
| *BCL11A* | intron | c.386-18893T>G | rs6545816 |
| *BCL11A* | intron | c.386-22075A>C | rs1427407 |
| *BCL11A* | intron | c.386-22379G>A | rs7599488 |

| | | | |
|---|---|---|---|
| *BCL11A* | intron | c.386-24002G>T | rs766432 |
| *BCL11A* | intron | c.386-24983T>C | rs4671393 |
| *BCL11A* | intron | c.386-33326C>T | rs10184550 |
| *HBS1L* | promoter | c.-380A>C | rs28384513 |

HGVS: Human Genome Variation Society; BCL11A: B-cell lymphoma/leukemia 11A; HBS1L: HBS1 Like Translational GTPase.

#### 6.4 No abnormal blood cell proliferation was observed in the bone marrow of F001 and F002

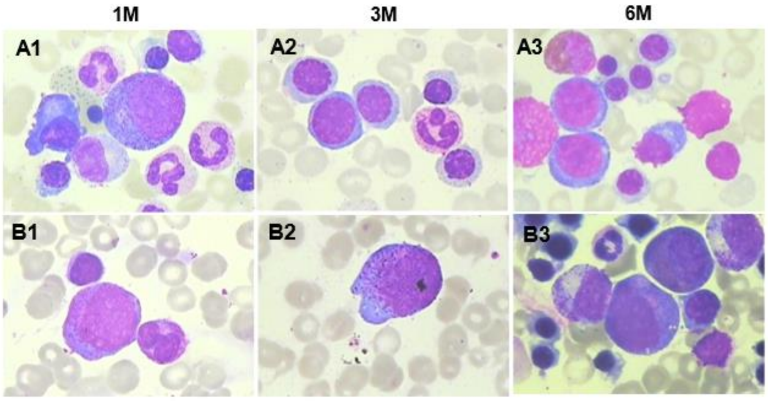


**Figure S6. The images of bone marrow smear of F001 and F002 at the 1, 3 and 6 months after gene therapy.** The upper panel represents F001, and A1, A2 and A3 represent 1 month, 3 months and 6 months after gene therapy respectively. The lower panel B represents F002, and B1, B2 and B3 represent 1 month, 3 months and 6 months after gene therapy respectively.

#### 6.5 Adverse events (AEs)

There were totally 27 AEs in subjects F001 and F002 from the signing of the informed consent to the most recent visit. Table S3 displays the various of AEs of F001 and F002 until last follow-up visit.

**Table S 3-1 AEs in F001 and F002 patients during myeloablative conditioning**

| Name of AEs | Grade 1/2 | Grade 3/4 | Total number |
|---|---|---|---|
| Nausea | 2 | 0 | 2 |
| Abdominal pain | 1 | 0 | 1 |
| Gingival infection | 1 | 0 | 1 |
| Total number | 4 | 0 | 4 |

**Table S 3-2 AEs in F001 and F002 patients in the transplant unit**

| Name of AEs | Grade 1/2 | Grade 3/4 | Total number |
|---|---|---|---|
| Hepatic VOD | 1 | 0 | 1 |
| Diarrhea | 1 | 0 | 1 |
| Oral mucositis | 2 | 0 | 2 |
| Febrile neutropenia | 0 | 2 | 2 |
| Neutropenia | 0 | 2 | 2 |
| Leukopenia | 0 | 2 | 2 |
| Thrombocytopenia | 0 | 2 | 2 |
| Hypokalemia | 1 | 0 | 1 |
| Vaginal bleeding | 1 | 0 | 1 |
| DH | 1 | 0 | 1 |
| Epistaxis | 1 | 0 | 1 |
| Pharyngalgia | 0 | 1 | 1 |
| Giddiness | 1 | 0 | 1 |
| Oral thrush | 0 | 1 | 0 |
| ES | 1 | 0 | 1 |
| Candida enteritis | 1 | 0 | 1 |
| Total number | 11 | 10 | 21 |

**Table S 3-3 AEs in F001 and F002 patients after discharge from transplant unit**

| Name of AEs | Grade 1/2 | Grade 3/4 | Total number |
|---|---|---|---|
| Hepatic VOD | 1 | 0 | 1 |
| Upper RTI | 1 | 0 | 1 |
| Total number | 2 | 0 | 2 |

Note: VOD: veno-occlusive disease; DH: dermatitis herpetiformis; ES: engraftment syndrome; RTI: respiratory tract infection

**6.6 The rising levels of soluble transferrin receptor (sTfR) and erythropoietin (EPO) indicates that F001 patient still has ineffective erythropoiesis**

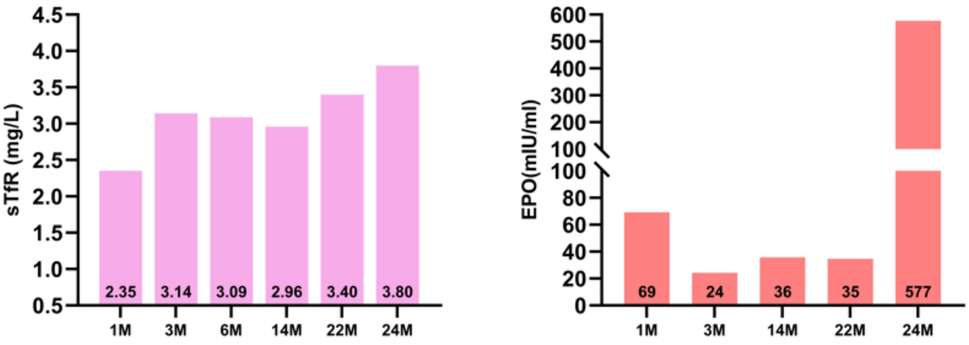


**Figure S7 The higher levels of sTfR and EPO have been observed after gene therapy in F001 patients.** It is indicated that the hemolysis status of F001 and F002 has been relived compared with prior to HGI-001 injection treatment.
